# Impact of Dapsone (MDT/WHO) on hematological tests of leprosy patients: A retrospective cohort study

**DOI:** 10.1101/2025.11.26.25341059

**Authors:** Cláudio Mariano da Silva, Gustavo Sartori Albertino, Júnia Adriano Wiezel, Fabiana Aparecida Correa Cinto, Fernanda André Martins da Cruz Perecin, Helena Barbosa Lugão, Filipe Rocha Lima, Marco Andrey Cipriani Frade

## Abstract

**Background:** Dapsone, a component of the WHO-recommended multidrug therapy (MDT/WHO) for leprosy, can induce oxidative hemolysis through its hydroxylamine metabolites. However, treatment guidelines lack objective laboratory thresholds to guide dapsone initiation. This study aimed to quantify the hematologic toxicity and evaluate potential liver effects of of dapsone and identify sex-specific cutoff values that predict anemia development.

**Methodology:** We conducted a retrospective cohort study using data from 258 patients treated for leprosy at a Brazilian tertiary hospital. Hematologic parameters were evaluated at diagnosis and 1, 3, 6, and 12 months of treatment. Patients were grouped by regimen: MDT/WHO (continuous dapsone use) and MDT/SUBSTITUTIVE (dapsone replaced after 1st month). ROC curve analyses were performed to identify optimal thresholds for predicting anemia by dapsone.

**Results/Principal Findings:** After the first month, patients had a mean Hb value decline of 1.14g/dL for MDT/WHO, and 1.65g/dL for MDT/SUBSTITUTIVE. Anemia percentual decreased by 58.5% in the MDT/SUBSTITUTIVE while only 13.0% in the MDT/WHO on month 12. For males, ROC analysis identified Hb<14.15g/dL (AUC 0.710), HCT<43.05% (AUC 0.711), and RBC<4.88×10⁶/mm³ (AUC 0.599) as optimal thresholds. For females, the cutoffs were Hb<13.65g/dL (AUC 0.767), HCT<38.85% (AUC 0.561), and RBC<4.455×10⁶/mm³ (AUC 0.590). Aspartate aminotransferase, alanine aminotransferase, and gamma-glutamyl transferase were also assessed; no clinically significant hepatotoxicity was found, nor correlation with Hb changes.

**Conclusions:** Dapsone induces early and sustained anemia during MDT/WHO. Our findings define sex-specific baseline thresholds for safer dapsone prescription. For males, absolute contraindication occurs with Hb<13.65g/dL (RR: 2.76[95%CI 1.92-3.98]), HCT<40.95%, or RBC<4.62×10⁶/mm³; relative contraindication for Hb<14.15g/dL (RR: 2.53[1.68-3.80]), HCT<43.05%, or RBC<4.88×10⁶/mm³. For females, absolute contraindication is with Hb<12.95g/dL (RR: 2.29[1.59-3.30]), HCT<36.5%, or RBC<4.2×10⁶/mm³; relative contraindication for Hb<13.65g/dL (RR: 2.64[1.41-4.93]), HCT<38.85%, or RBC<4.455×10⁶/mm³. These thresholds support individualized, laboratory-based eligibility criteria and early dapsone substitution in high-risk individuals.

**Author summary:** Leprosy remains an important public health challenge, particularly in countries where it is still endemic, and its treatment depends on a combination of drugs that includes dapsone. While dapsone is a key part of therapy, it often causes anemia, a condition where the blood cannot carry enough oxygen to meet the body’s needs. This side effect not only weakens patients but can also reduce their ability to complete treatment successfully. Until now, there were no clear laboratory values to help doctors decide when dapsone could be used safely or when it should be replaced, leaving both patients and health professionals uncertain. In this study, we followed people with leprosy treated at a hospital in Brazil and reviewed their blood tests at several points during one year of treatment. We found that anemia appeared quickly after therapy began, especially among women, and in many patients, it persisted until the end of treatment. Those who stopped dapsone early showed faster recovery. By analyzing blood counts before treatment, we identified values that predicted who was most at risk. Our findings provide practical thresholds that can support safer, more personalized treatment of leprosy, particularly in resource-limited settings where close monitoring is difficult.

## Introduction

Leprosy is an infectious disease with chronic progression, caused by the etiological agent Mycobacterium leprae. This bacillus affects the skin and peripheral nerves and can result in severe sequelae if early intervention is not implemented[1, 2]. The disease remains endemic in more than 140 countries worldwide[3]. Modes of transmission include household contact with infected individuals, exposure to oral and nasal secretions from untreated patients. Household contacts account for up to 28% of new cases and have a 4 to 10-fold higher risk of long-term infection[4, 5].

The availability of free medication for the treatment of leprosy has significantly improved cure rates and reduced the number of new cases. However, the disease remains a major cause of morbidity, particularly among socially vulnerable populations. In 2023, a total of 182,815 new cases of leprosy were reported across 184 countries, areas, and territories, with 39.8% (72,845) of cases occurring in women and 5.6% (10,322) in children[6]. Brazil maintained its position among the countries with the highest detection rates, reporting 22,773 new diagnoses – second only to India[7]. Leprosy remains the leading cause of infectious neuropathy in tropical and subtropical regions, potentially resulting in motor and sensory impairments, as well as limb deformities[8].

To control transmission and achieve microbiological cure, the World Health Organization (WHO) introduced multidrug therapy (MDT/WHO) in the 1980s, combining rifampicin, clofazimine, and dapsone. The WHO Guidelines Development Group recommends a uniform 3-drug regimen, which contains rifampicin, dapsone, and clofazimine for all leprosy patients, with a 6-month course for paucibacillary and 12 months for multibacillary disease. This approach is supported by evidence favoring superior outcomes of 3-drug therapy in paucibacillary cases compared to 2-drug regimens. Importantly, it mitigates risks of misclassification between disease forms and offers implementation advantages by standardizing treatment across both classifications[9]. In cases of intolerance or contraindication, ofloxacin and/or minocycline may be used as alternatives, or the ROM (Rifampicin, Ofloxacin and Minocycline) regimen or RIMOXCLAMIN (Rifampicin, Minocycline, Ofloxacin, Clofazimine, and Moxifloxacin) may be employed in specific scenarios [10, 11].

Dapsone (4,4’-diaminodiphenylsulfone) is an aromatic amine from the sulfone family, synthesized in 1945 from p-chloronitrobenzene. Its antibacterial action results from structural similarity to sulfonamides, acting as a competitive inhibitor of the enzyme dihydropteroate synthase (DHPS), which blocks folate synthesis and impairs DNA replication[12]. However, despite its therapeutic efficacy, dapsone is associated with hematologic toxicity, notably dose-dependent hemolytic anemia and methemoglobinemia. These reactions are mainly attributed to its N-hydroxylated metabolites, especially dapsone hydroxylamine, produced by cytochrome P450 enzymes (CYP2C9 and CYP2C19)[13–16].

These metabolites increase oxidative stress, promote reactive oxygen species (ROS) formation, and damage erythrocytes by generating methemoglobin. Individuals with glucose-6-phosphate dehydrogenase (G6PD) deficiency are especially vulnerable due to reduced enzymatic capacity to neutralize oxidative insults, making dapsone contraindicated in this population[12, 17]. Yet, even in patients with normal G6PD activity, the hematologic effects of dapsone – such as declines in hemoglobin (Hb) and hematocrit (HCT) – are well documented but insufficiently quantified in large, real-world populations.

Despite its long-standing use, few studies have systematically evaluated hematologic changes throughout the course of MDT/WHO, particularly with standardized follow-up points and comparative analyses. Moreover, current treatment guidelines do not define a minimum Hb threshold for safe dapsone initiation, posing a challenge for clinicians facing patients with borderline anemia or underlying vulnerabilities.

Notably, current treatment guidelines do not provide objective laboratory thresholds to guide dapsone initiation, particularly in patients with borderline hematologic profiles. Our findings aim to contribute to discussions on clinical safety criteria for initiating MDT/WHO and optimizing treatment protocols in endemic areas.

## Material and methods

### Study design and objectives

#### Data sources and collection

Data were obtained from electronic medical records and laboratory databases maintained by the Information Center at HC FMRP-USP. Variables extracted included demographic information, treatment regimens, and laboratory parameters. Only patients with complete hematological data for all five follow-up periods were included.

Additionally, glucose-6-phosphate dehydrogenase (G6PD) enzyme activity was assessed in a subset of patients, according to clinical discretion and availability of the test.

#### Data treatment strategy

Following the acquisition of raw data from the spreadsheet provided by the Information Center of HC FMRP-USP, filtering steps were implemented to ensure the quality, integrity and reliability of the data to be analyzed.

The sample in this study consisted of 46,468 sets of examination records from 3,641 patients. Each set of records represents a specific patient along with the respective information on hematimetric examinations conducted during the leprosy treatment follow-up. Figure 1 presents the decision pathway used to determine data eligibility for this study.

**Figure 1.**
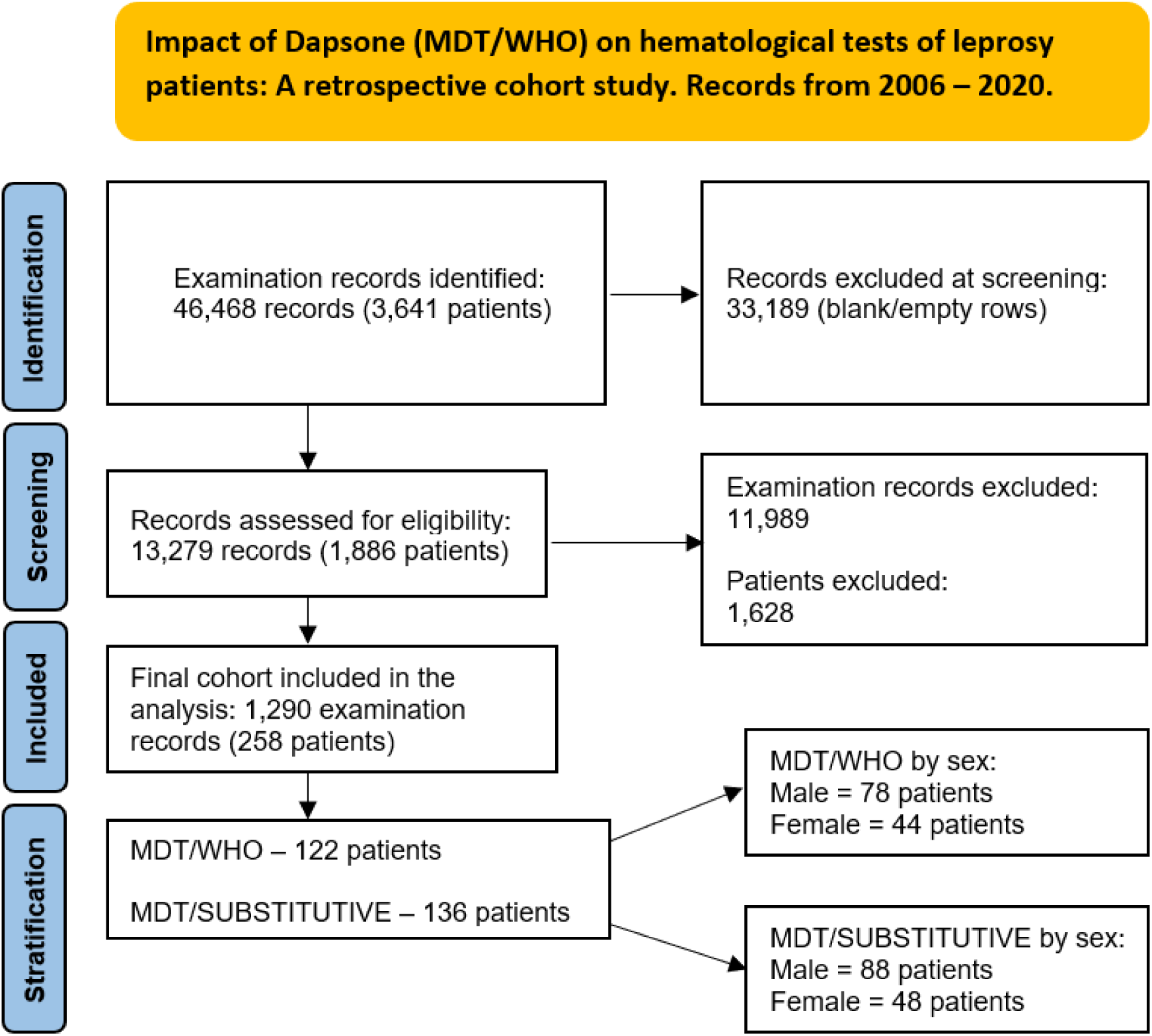
Flow diagram of data selection and exclusion process. From a total of 46,468 examination records (3,641 patients) identified between 2006 and 2020, 33,189 blank or empty records were excluded. After eligibility assessment of 13,279 records (1,886 patients), 11,989 examination records and 1,628 patients were excluded. The final cohort included 1,290 valid examination records from 258 patients, divided into two groups: MDT/WHO (n=122) and MDT/SUBSTITUTIVE (n=136). Each group was further stratified by sex.

Initially, all rows containing blank data related to hematimetric indices were identified and removed. Consequently, 13,279 rows of laboratory examination data from 1,886 patients were retained, forming the initial dataset for analysis. The next step was to select the data related to the periods of interest defined in the Follow-Up Measures. Patients who did not have corresponding examinations for all 5 periods were excluded from the analysis. This step was undertaken to maintain uniform sample sizes across each designated time interval and facilitate a more accurate temporal correlation between cause and effect.

After data treatment, we retained 1,290 rows of examination data from 258 patients. This final sample includes detailed information for all five periods of interest for each patient, enabling a more robust and precise analysis of hematimetric indices during leprosy treatment. With the filters applied, the analysis period spans from 2006 to 2020, with no eligible patients from 2001 to 2005.

#### Treatment protocol and group allocation

All 258 patients initially began treatment with the standard MDT/WHO regimen, which includes dapsone as one of its core components. After the first month, some patients developed symptomatic anemia by the use of dapsone, and then it was replaced by either ofloxacin or minocycline, depending on clinical indication and drug availability, resulting in a modified therapeutic regimen. This group was designated as MDT/SUBSTITUTIVE. The patients who kept the standard MDT regimen till the end of treatment were constituting the MDT/WHO group for comparative analysis.

#### Stratification by sex, therapeutic regimen, and anemia severity

In this study, anemia was defined and graded according to the *2024 WHO Guideline on Hemoglobin Cutoffs to Define Anaemia in Individuals and Populations[18]*. These updated thresholds were established based on the 5th percentiles of Hb distributions in healthy, ethnically diverse individuals from high-income countries, including Australia, China, Europe, North America, and South America.

For adult populations, we applied the recommended cutoffs for men aged 15–65 years and non-pregnant women aged 15–65 years. Anemia was classified into four categories:

● **No anemia:** Hb ≥13.0g/dL (men), Hb ≥12.0g/dL (women)
● **Mild anemia**: Hb 11.0–12.9g/dL (men), Hb 11.0–11.9g/dL (women)
● **Moderate anemia**: Hb 8.0–10.9g/dL (both sexes)
● **Severe anemia**: Hb<8.0g/dL (both sexes)

These categories were used to stratify anemia severity in the analysis of Hb trends and treatment-related hematological toxicity.

### Data analysis

For the analysis and statistical treatment of the obtained data, GraphPad Prism v.10 software (GraphPad Software, San Diego, CA) was used. The assessment of data normality was conducted considering the sample size and applying both the D’Agostino-Pearson and Shapiro-Wilk tests to classify sample distributions as parametric or non-parametric. Once classified as non-parametric, the Mann-Whitney or Kruskal-Wallis tests were used for group comparisons, while the t-test or One Way Analysis of Variance (ANOVA) was used for parametric samples. The significance level was set at 95% (alpha=0.05), with a p-value less than or equal to 0.05 considered statistically significant.

Diagnostic performance indicators were computed using standard 2×2 contingency tables (TP=true positive; TN=true negative; FP=false positive; FN=false negative).[19, 20] Sensitivity was calculated as TP/(TP+FP), and specificity as TN/(TN+FN). Accuracy was defined as (TP+TN)/(TP+TN+FP+FN). Positive predictive value (PPV) and negative predictive value (NPV) were obtained as TP/(TP+FN) and TN/(TN+FP), respectively. Relative risk (RR) was estimated as [TP/(TP+FN)]/[FP/(FP+TN)] [21, 22]. From these metrics we derived the Youden Index (J=Sensitivity+Specificity–1)[23] and the Sensitivity×Specificity product to identify optimal cutoffs. These computations followed current recommendations for diagnostic test evaluation.

Statistical power analysis was conducted using G*Power software version 3.1.9.7 to compare two independent proportions through Fisher’s exact test (one-tailed). Based on expected anemia recovery rates between month 1 and month 12 of 20% in the MDT/WHO group and 50% in the MDT/SUBSTITUTIVE group, with a significance level of 5% (α=0.05) and desired power of 95% (1−β=0.95), the estimated minimum sample size required was 59 patients per group (total n=118). Given the actual study sample (n=258; MDT/WHO=122; MDT/SUBSTITUTIVE=136), the resulting statistical power was 95.2%, confirming that the available sample size was adequate to detect clinically meaningful differences between treatment regimens.

ROC (Receiver Operating Characteristic) curve analyses and associated graphical representations were performed using R software version 4.5.0 (The R Foundation for Statistical Computing, Vienna, Austria). Packages such as pROC and ggplot2 were employed to calculate diagnostic performance metrics, plot the ROC curves, and identify optimal cutoffs using the Youden Index.

### Ethics statement

According to the Helsinki Declaration, this retrospective study was approved by the Research Ethics Committee at the Clinics Hospital of Ribeirão Preto Medical School, University of São Paulo (protocol n° 4.102.290-06/22/2020). All analyzed data are anonymous and protected in accordance with privacy and research ethics standards.

## Results

### Demographic profile of the study population

The demographic characteristics of the patients are described in Table 1. The study sample comprised 258 individuals undergoing treatment for leprosy. Among them, 64.3% are male (n=166) and 35.7% female (n=92). The mean age was 52 (SD=15) years, reflecting a predominance of adults in middle age. With regard to ethnicity, 74.4% self-identified as white, followed by 16.3% brown/mixed-race and 9.3% Black individuals. Most patients were residents of the state of São Paulo (SP) (96.1%, n=248), with a minority from Minas Gerais (MG) (3.1%, n=8), and isolated cases from Goiás (GO) and Tocantins (TO) (each with 0.4%, n=1).

**Table 1.** Demographic data of the patients (N = 258). Sample size (n) and its percentage (%) in relation to sex, ethnicity, federative unit of origin.

| Characteristic |  | Patients (N=258) |  |
| --- | --- | --- | --- |
|  |  | n | % |
| Sex | Male | 166 | 64.3 |
|  | Female | 92 | 35.7 |
| Age (years) | Mean (SD) | 52 | (15) |
| Ethnicity | White | 192 | 74.4 |
|  | Brown (Mixed-race) | 42 | 16.3 |
|  | Black | 24 | 9.3 |
| Federative unit of origin | SP (Southeast) | 248 | 96.1 |
|  | MG (Southeast) | 8 | 3.1 |
|  | GO (Central-West) | 1 | 0.4 |
|  | TO (North) | 1 | 0.4 |

SD: Standard deviation. SP: São Paulo; MG: Minas Gerais; GO: Goiás; TO: Tocantins.

### G6PD testing

Among the 144 patients with G6PD testing available, 143 exhibited normal enzymatic activity and only 1 case of G6PD deficiency was detected. In the remaining 114 patients, the test was not performed. This distribution suggests that the hematological alterations observed in this study are unlikely to be attributable to G6PD deficiency and rather reflect the intrinsic oxidative toxicity of dapsone in individuals with preserved enzymatic function.

### Temporal evolution in hematological parameters during MDT/WHO therapy

The hematological parameters across the treatment period are summarized in Table 2. The mean red blood cell (RBC) count exhibited a decline after the initiation of treatment, dropping from 4.6×10⁶/mm³ at baseline to 4.1×10⁶/mm³ in the first month. These values remained stable at the third and sixth months, followed by a partial recovery to 4.4×10⁶/mm³ by the twelfth month. Despite this increase, RBC counts did not fully return to baseline levels.

**Table 2.** Hematological indices measured during leprosy treatment (N = 258), for red blood cell count, hemoglobin, and hematocrit across five time points.

| Component |  | M0 | M1 | M3 | M6 | M12 |
| --- | --- | --- | --- | --- | --- | --- |
| Hemoglobin (g/dL) | Mean | 13.7 | 12.3 | 12.5 | 12.7 | 13.2 |
|  | SD | 1.69 | 1.89 | 1.68 | 1.60 | 1.71 |
|  | Lower 95% CI | 13.5 | 12.0 | 12.3 | 12.5 | 13.0 |
|  | Upper 95% CI | 13.9 | 12.5 | 12.7 | 12.9 | 13.4 |
|  | CV (%) | 12.4 | 15.4 | 13.5 | 12.6 | 13.0 |
| Hematocrit (%) | Mean | 41 | 37 | 38 | 39 | 40 |
|  | SD | 4.8 | 5.6 | 4.9 | 4.6 | 5.0 |
|  | Lower 95% CI | 40 | 36 | 37 | 38 | 39 |
|  | Upper 95% CI | 42 | 38 | 39 | 39 | 40 |
|  | CV (%) | 12 | 15 | 13 | 12 | 13 |
| Red Blood Cells (10<sup>6</sup>cells/mm<sup>3</sup>) | Mean | 4.6 | 4.1 | 4.1 | 4.2 | 4.4 |
|  | SD | 0.58 | 0.67 | 0.63 | 0.61 | 0.64 |
|  | Lower 95% CI | 4.51 | 4.00 | 4.03 | 4.14 | 4.28 |
|  | Upper 95% CI | 4.66 | 4.17 | 4.19 | 4.29 | 4.44 |
|  | CV (%) | 12.7 | 16.3 | 15.3 | 14.4 | 14.6 |

SD: Standard deviation; CI: Confidence interval; CV: Coefficient of variation. Time points are denoted as M0 (baseline), M1 = month 1, M3 = month 3, M6 = month 6, M12 = month 12.

Hb concentrations followed a similar trend. A marked reduction was observed after the first month of therapy (from 13.7g/dL to 12.3g/dL), with gradual increases noted at subsequent follow-ups, culminating at 13.2g/dL in the twelfth month. However, final values remained slightly lower than the pre-treatment mean. The standard deviation and coefficient of variation were highest in the early treatment phase, indicating greater variability in Hb response.

Regarding HCT values, the initial mean of 41% decreased to 37% after the first month, with a progressive recovery observed throughout the follow-up, reaching 40% by month twelve. Although the HCT trajectory mirrored that of Hb, the amplitude of change was relatively lower.

Across all three indices, the first month of treatment marked the period of greatest hematological impact, followed by progressive improvement. However, none of the parameters fully reverted to baseline levels by the end of the 12-month period.

After the first month of therapy, out of the 258 patients, 136 (52.7%) developed adverse clinical effects, primarily symptoms consistent with drug-induced anemia, and they were unable to continue dapsone use. Therefore, these patients were included in the MDT/SUBSTITUTIVE regimen. The remaining 122 patients (47.3%) tolerated dapsone and completed the full 12-month course with the unaltered MDT/WHO regimen.

The evolution of Hb levels and the proportion of anemic patients across the treatment period is detailed in Table 3. At baseline (M0), patients in the MDT/WHO group exhibited a higher mean Hb concentration (14.13g/dL) compared to those in the MDT/SUBSTITUTIVE group (13.28g/dL). Following initiation of treatment, both groups showed a decline in Hb values by the first month (M1), with the reduction more pronounced in the MDT/SUBSTITUTIVE group (−1.65g/dL vs. −1.14g/dL). Figure 2 illustrates the evolution of Hb, HCT and RBC for the MDT/WHO and MDT/SUBSTITUTIVE regimens.

**Figure 2.**
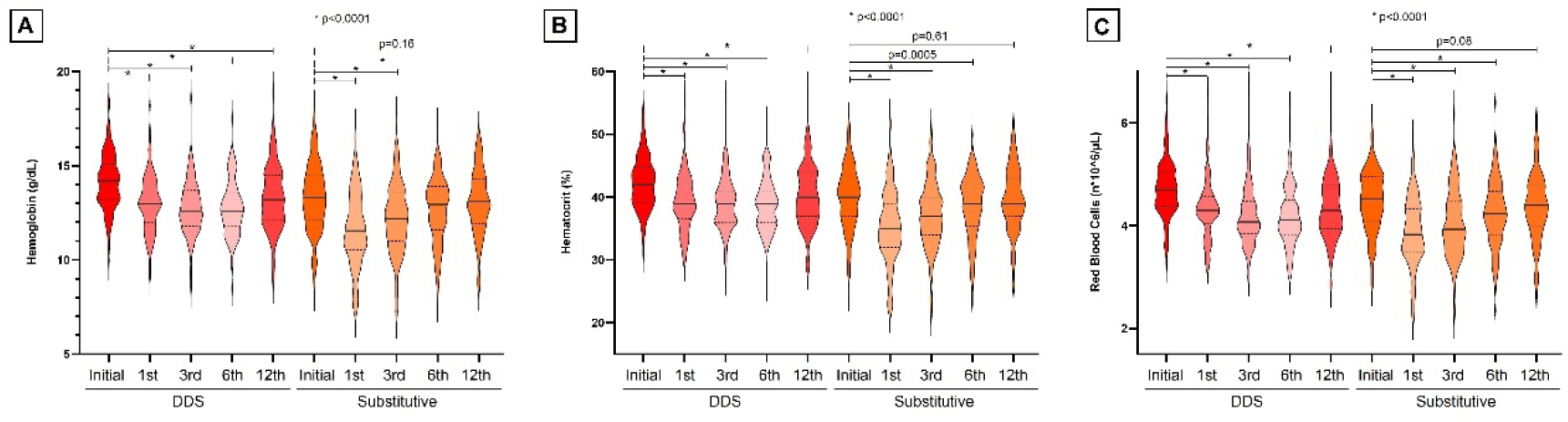
Temporal evolution of hematological parameters during leprosy treatment, according to therapeutic regimen. (A) Mean hemoglobin values (g/dL). (B) Mean hematocrit values (%). (C) Mean Red Blood Cells values (×10⁶/mm³). Statistical comparisons were performed using paired t-tests (vs. M0). Asterisks indicate p<0.0001.

**Table 3.** Evolution of hemoglobin levels and anemia prevalence during treatment, stratified by therapeutic regimen.

| Component |  | MDT/WHO (n=122) |  |  |  |  | MDT/SUBSTITUTIVE (n=136) |  |  |  |  |
| --- | --- | --- | --- | --- | --- | --- | --- | --- | --- | --- | --- |
|  |  | M0 | M1 | M3 | M6 | M12 | M0 | M1 | M3 | M6 | M12 |
| Anemic Samples | n | 15 | 46 | 51 | 58 | 40 | 30 | 82 | 61 | 43 | 34 |
|  | % | 12.3 | 37.7 | 41.8 | 47.5 | 32.8 | 22.1 | 60.3 | 44.9 | 31.6 | 25.0 |
|  | Δ (% compared to M0) | — | 206.7 | 240 | 286.7 | 166.7 | — | 173.3 | 103.3 | 43.3 | 13.3 |
| Hemoglobin (g/dL) | Mean | 14.13 | 12.99 | 12.76 | 12.75 | 13.3 | 13.28 | 11.63 | 12.19 | 12.63 | 13.06 |
|  | SD | 1.49 | 1.52 | 1.43 | 1.46 | 1.72 | 1.77 | 1.97 | 1.84 | 1.7 | 1.71 |
|  | p-value (compared to M0) | — | * | * | * | * | * | * | * | * | 0.1582 |
| Hematocrit (%) | Mean | 42.25 | 39.24 | 38.90 | 38.70 | 40.17 | 39.85 | 35.33 | 37.11 | 38.38 | 39.61 |
|  | SD | 4.18 | 4.50 | 4.19 | 4.14 | 4.91 | 5.05 | 5.82 | 5.34 | 5.00 | 5.14 |
|  | p-value (compared to M0) | — | * | * | * | * | * | * | * | 0.0005 | 0.6112 |
| Red Blood Cells (10<sup>6</sup>cells/mm<sup>3</sup>) | Mean | 4.72 | 4.32 | 4.19 | 4.18 | 4.35 | 4.47 | 3.88 | 4.04 | 4.24 | 4.37 |
|  | SD | 0.54 | 0.58 | 0.57 | 0.57 | 0.61 | 0.59 | 0.67 | 0.67 | 0.64 | 0.66 |
|  | p-value (compared to M0) | — | * | * | * | * | * | * | * | * | 0.0793 |

Mean hemoglobin values (g/dL), standard deviations (SD), and the number and percentage of anemic patients during treatment with MDT/WHO or MDT/SUBSTITUTIVE regimens. Time points: M0 = baseline, M1 = month 1, M3 = month 3, M6 = month 6, M12 = month 12. Statistical comparisons (paired t-tests) were performed between M0 and each subsequent time point. \*Asterisks denote p<0.0001. Dashes (—) indicate null value.

In the MDT/WHO group, mean Hb levels remained low until M6 and rose by M12 (13.3g/dL), though still below baseline. The percentage of anemic patients increased from 13.1% at M0 to 43.4% at M3, with a slight decline to 36.1% by M12. All comparisons between M0 and subsequent time points (M1, M3, M6, M12 were statistically different (p<0.0001).

#### Effect of dapsone replacement in MDT

In contrast, the MDT/SUBSTITUTIVE group, which discontinued dapsone after M1, demonstrated an initial worsening in anemia frequence (66.9% at M1), followed by progressive improvement over time, reaching 34.6% at M12. Hb levels approached baseline by M12 (13.06g/dL), and the difference between M0 and M12 was not statistically different (p=0.1582).

The sex-stratified evolution of Hb levels and anemia frequence in patients under the MDT/WHO regimen is presented in Figure 3 and detailed in Table 4. At baseline (M0), male patients exhibited a higher mean Hb value (14.44g/dL) as compared to females (13.57g/dL). Both groups experienced a decline in Hb by month 1: a reduction of 1.12g/dL in males and 1.16g/dL in females.

**Figure 3.**
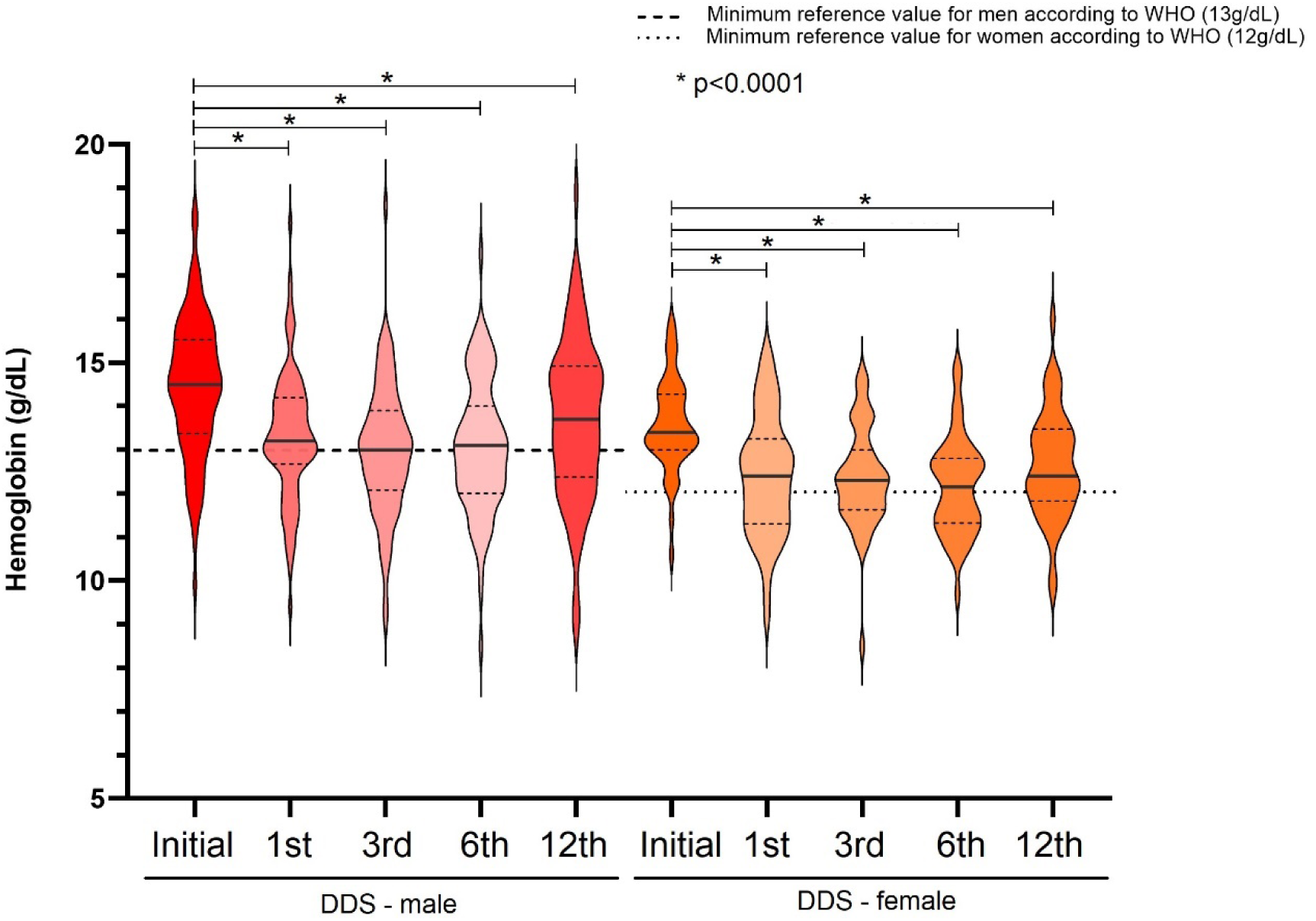
Temporal evolution of hemoglobin levels and anemia prevalence in patients under MDT/WHO treatment, stratified by sex. Mean hemoglobin values (g/dL) and proportion of anemic patients at five time points (M0, M1, M3, M6, M12) in male (n=78) and female (n=44) patients. Both sexes exhibited a decline in hemoglobin after the first month of treatment, followed by partial recovery. The percentage of anemic patients increased in both groups, peaking at month 6 and decreasing by month 12. All comparisons with baseline values were statistically significant (p<0.0001). Error bars represent standard deviation.

**Table 4.** Evolution of hemoglobin levels and anemia prevalence during treatment with MDT/WHO, stratified by male and female.

| HEMOGLOBIN |  | MDT/WHO (n=122) |  |  |  |  |  |  |  |  |  |
| --- | --- | --- | --- | --- | --- | --- | --- | --- | --- | --- | --- |
|  |  | Male (n=78) |  |  |  |  | Female (n=44) |  |  |  |  |
|  |  | M0 | M1 | M3 | M6 | M12 | M0 | M1 | M3 | M6 | M12 |
| Mean |  | 14.44 | 13.32 | 12.99 | 13.07 | 13.66 | 13.57 | 12.41 | 12.35 | 12.17 | 12.65 |
| SD |  | 1.60 | 1.50 | 1.52 | 1.54 | 1.85 | 1.08 | 1.38 | 1.16 | 1.11 | 1.24 |
| Anemic Samples | n | 13 | 29 | 35 | 38 | 27 | 2 | 17 | 16 | 20 | 13 |
|  | % | 16.7 | 37.2 | 44.9 | 48.7 | 34.6 | 4.5 | 38.6 | 36.4 | 45.5 | 29.5 |
| p-value (compared to M0) |  | — | * | * | * | * | — | * | * | * | * |

Mean hemoglobin values (g/dL), standard deviations (SD), and the number and percentage of anemic patients during treatment with MDT/WHO, stratified by sex. Time points: M0 = baseline, M1 = month 1, M3 = month 3, M6 = month 6, M12 = month 12. Statistical comparisons (paired t-tests) were performed between M0 and each subsequent time point. \*Asterisks denote $p < 0.0001$ . Dashes (—) indicate null value.

Among men, Hb levels stabilized around months 3 and 6, with gradual recovery by month 12 (13.66g/dL). Anemia frequence in this group rose substantially from 16.7% at baseline to 48.7% at month 6, subsequently declining to 34.6% at month 12. All time points showed statistically significant differences compared to baseline (p<0.0001).

For female patients, Hb reached the lowest mean at month 6 (12.17g/dL). The frequence of anemia increased from 4.5% at baseline to 45.5% on month 6 and declined to 29.5% by month 12. Despite smaller variations in mean Hb, the relative increase in anemia prevalence was substantial in women.

The sex-stratified evolution of Hb levels and anemia prevalence among patients treated with the MDT/SUBSTITUTIVE regimen is presented in Figure 4 and detailed in Table 5. At baseline, male patients exhibited higher mean Hb values (13.53g/dL) than female patients (12.83g/dL). Both groups showed a sharp decline after the first month of treatment, reaching 11.88g/dL in males and 11.17g/dL in females.

**Figure 4.**
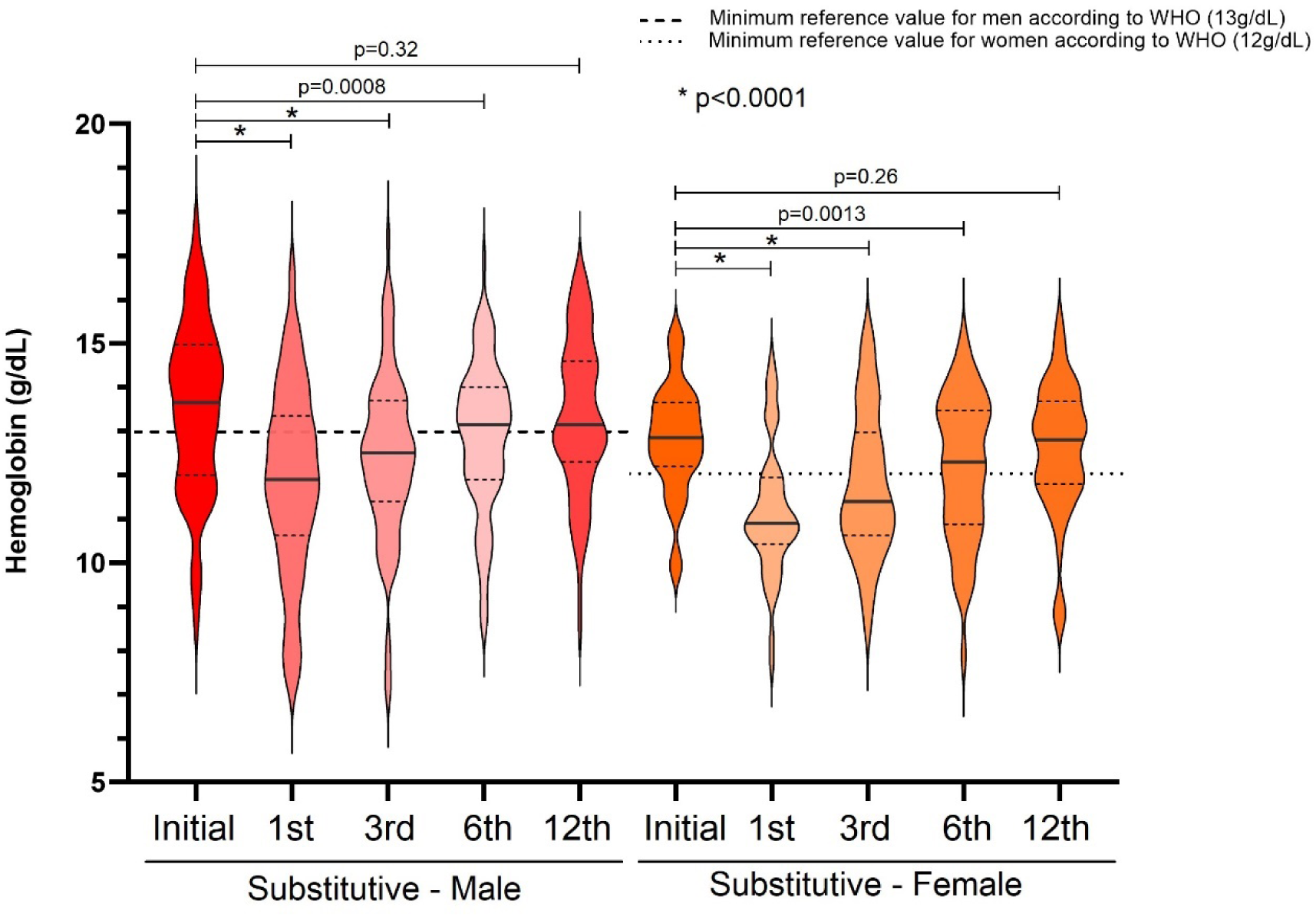
Temporal evolution of hemoglobin levels and anemia prevalence in patients under the MDT/SUBSTITUTIVE regimen, stratified by sex. Mean hemoglobin values (g/dL) and proportion of anemic patients at five time points (M0, M1, M3, M6, M12) in male (n=88) and female (n=48) patients. Both sexes exhibited a decline in hemoglobin after the first month of treatment, followed by almost complete recovery. The percentage of anemic patients increased in both groups, peaking at month 1 and decreasing after the dapsone substitution. The comparisons with baseline values for the months 1, 3 and 6 were statistically significant. There was no statistically significant difference between month 12 and the baseline values. Error bars represent standard deviation.

**Table 5.** Evolution of hemoglobin levels and anemia prevalence during treatment with MDT/SUBSTITUTIVE, stratified by male and female.

| HEMOGLOBIN |  | MDT/SUBSTITUTIVE (n=136) |  |  |  |  |  |  |  |  |  |
| --- | --- | --- | --- | --- | --- | --- | --- | --- | --- | --- | --- |
|  |  | Male (n=88) |  |  |  |  | Female (n=48) |  |  |  |  |
|  |  | M0 | M1 | M3 | M6 | M12 | M0 | M1 | M3 | M6 | M12 |
| Mean |  | 13.53 | 11.88 | 12.42 | 12.89 | 13.33 | 12.83 | 11.17 | 11.78 | 12.14 | 12.59 |
| SD |  | 1.962 | 2.159 | 1.912 | 1.701 | 1.77 | 1.249 | 1.483 | 1.646 | 1.636 | 1.493 |
| Anemic Samples | n | 33 | 61 | 55 | 39 | 39 | 9 | 36 | 28 | 20 | 16 |
|  | % | 37.5 | 69.3 | 62.5 | 44.3 | 44.3 | 18.8 | 75.0 | 53.8 | 41.7 | 33.3 |
| p-value (compared to M0) |  | – | * | * | 0.0008 | 0.32 | – | * | * | 0.0013 | 0.26 |

Mean hemoglobin values (g/dL), standard deviations (SD), and the number and percentage of anemic patients during treatment with MDT/SUBSTITUTIVE, stratified by sex. Time points: M0 = baseline, M1 = month 1, M3 = month 3, M6 = month 6, M12 = month 12. Statistical comparisons (paired t-tests) were performed between M0 and each subsequent time point. \*Asterisks denote $p < 0.0001$ . Dashes (–) indicate null value.

In males, the frequence of anemia increased from 37.5% at baseline to 69.3% at month 1, then decreased progressively, reaching 44.3% at month 12. Hb levels returned to values close to baseline (13.33g/dL), with no statistical differences between month 12 and baseline (p=0.32).

In females, anemia frequence increased from 18.8% at baseline to 75.0% at month 1. On month 12, 33.3% of participants remained anemic, with a mean Hb level of 12.59g/dL - statistically indistinguishable from baseline (p=0.26).

Percentage of patients classified by anemia severity level (no anemia, mild, moderate, severe) were stratified by sex and therapeutic group, and Table 6 shows this set of data. Among male patients treated with the MDT/WHO regimen, there was a progressive decline in the proportion without anemia, from 83.3% at baseline to 51.3% at month 6, followed by partial recovery to 65.4% at month 12 (M12). The percentage of men with mild anemia increased to a peak of 43.6% at M6, while moderate anemia remained relatively low throughout the follow-up (maximum of 9.0% at M3). No cases of severe anemia were observed in this group.

**Table 6.**
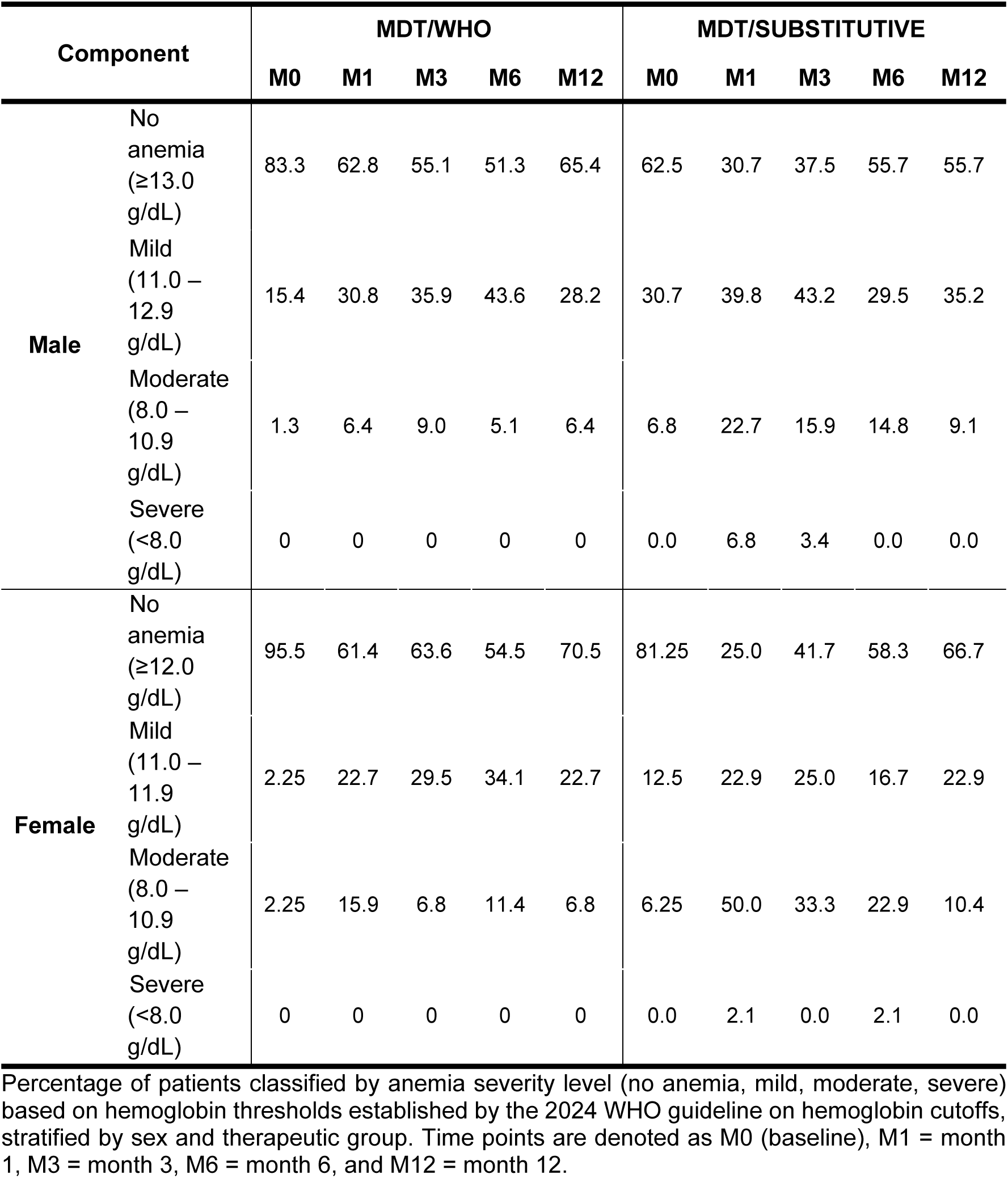
Temporal distribution of anemia severity by sex and therapeutic regimen (MDT/WHO vs. MDT/SUBSTITUTIVE).

| Component |  | MDT/WHO |  |  |  |  | MDT/SUBSTITUTIVE |  |  |  |  |
| --- | --- | --- | --- | --- | --- | --- | --- | --- | --- | --- | --- |
|  |  | M0 | M1 | M3 | M6 | M12 | M0 | M1 | M3 | M6 | M12 |
| **Male** | No anemia (≥13.0 g/dL) | 83.3 | 62.8 | 55.1 | 51.3 | 65.4 | 62.5 | 30.7 | 37.5 | 55.7 | 55.7 |
|  | Mild (11.0 – 12.9 g/dL) | 15.4 | 30.8 | 35.9 | 43.6 | 28.2 | 30.7 | 39.8 | 43.2 | 29.5 | 35.2 |
|  | Moderate (8.0 – 10.9 g/dL) | 1.3 | 6.4 | 9.0 | 5.1 | 6.4 | 6.8 | 22.7 | 15.9 | 14.8 | 9.1 |
|  | Severe (<8.0 g/dL) | 0 | 0 | 0 | 0 | 0 | 0.0 | 6.8 | 3.4 | 0.0 | 0.0 |
| **Female** | No anemia (≥12.0 g/dL) | 95.5 | 61.4 | 63.6 | 54.5 | 70.5 | 81.25 | 25.0 | 41.7 | 58.3 | 66.7 |
|  | Mild (11.0 – 11.9 g/dL) | 2.25 | 22.7 | 29.5 | 34.1 | 22.7 | 12.5 | 22.9 | 25.0 | 16.7 | 22.9 |
|  | Moderate (8.0 – 10.9 g/dL) | 2.25 | 15.9 | 6.8 | 11.4 | 6.8 | 6.25 | 50.0 | 33.3 | 22.9 | 10.4 |
|  | Severe (<8.0 g/dL) | 0 | 0 | 0 | 0 | 0 | 0.0 | 2.1 | 0.0 | 2.1 | 0.0 |

Percentage of patients classified by anemia severity level (no anemia, mild, moderate, severe) based on hemoglobin thresholds established by the 2024 WHO guideline on hemoglobin cutoffs, stratified by sex and therapeutic group. Time points are denoted as M0 (baseline), M1 = month 1, M3 = month 3, M6 = month 6, and M12 = month 12.

In contrast, males in the MDT/SUBSTITUTIVE group showed a more hematological response, with only 30.7% without anemia at M1. Moderate anemia reached a peak of 22.7% at M1, but progressively decreased over time, reaching 9.1% at M12, with complete resolution of severe anemia after M3.

Female patients presented more pronounced and sustained hematological alterations in both regimens. In the MDT/WHO group, the percentage without anemia dropped from 95.5% at baseline to 54.5% at M6, with mild and moderate anemia peaking at 34.1% and 11.4%, respectively. Similar to males, no cases of severe anemia were recorded.

Female patients under MDT/SUBSTITUTIVE therapy had a sharper decline in normal Hb levels, with only 25.0% without anemia at M1, and moderate anemia affecting 50.0% of the group at that same time point. Despite a progressive recovery, only 66.7% were without anemia at M12, and mild to moderate anemia persisted in one third of the group.

### Relative risk of anemia after dapsone initiation

Table 7 presents the stratification of anemia incidence at M1 among 120 initially non-anemic male patients, according to baseline Hb levels at M0. For patients with Hb≤13.0g/dL, the anemia percentual at M1 was 100.0%. This percentage remained above 80% for Hb values up to 13.6g/dL, with a relative risk (RR) ranging from 2.05 to 2.76.

**Table 7.** Relative risk of anemia after dapsone initiation in 120 non-anemic male and 81 non-anemic female patients, stratified by baseline hemoglobin level (M0).

| Hb cutoff level<br>(g/dL at M0) |  | ≤ cutoff |  |  | > cutoff |  |  | RR | 95% CI |  |
| --- | --- | --- | --- | --- | --- | --- | --- | --- | --- | --- |
|  |  | n (M0) | anemic (at M1) |  | n (M0) | anemic (at M1) |  |  | Upper | Lower |
|  |  |  | n | % |  | n | % |  |  |  |
| Male | 12.95 | — | — | — | 120 | 47 | 39.2 | — | — | — |
|  | 13.15 | 3 | 3 | 100.0 | 117 | 44 | 37.6 | 2.66 | 2.11 | 3.36 |
|  | 13.55 | 18 | 15 | 83.3 | 102 | 32 | 31.4 | 2.66 | 1.86 | 3.78 |
|  | 13.65 | 22 | 18 | 81.8 | 98 | 29 | 29.6 | 2.76 | 1.92 | 3.98 |
|  | 13.75 | 25 | 18 | 72.0 | 95 | 29 | 30.5 | 2.36 | 1.60 | 3.48 |
|  | 13.95 | 28 | 20 | 71.4 | 92 | 27 | 29.3 | 2.43 | 1.64 | 3.61 |
|  | 14.05 | 31 | 22 | 71.0 | 89 | 25 | 28.1 | 2.53 | 1.69 | 3.77 |
|  | 14.15 | 33 | 23 | 69.7 | 87 | 24 | 27.6 | 2.53 | 1.68 | 3.80 |
|  | 14.95 | 69 | 35 | 50.7 | 51 | 12 | 23.5 | 2.16 | 1.25 | 3.72 |
| Female | 11.95 | — | — | — | 81 | 42 | 51.9 | — | — | — |
|  | 12.15 | 2 | 2 | 100.0 | 79 | 40 | 50.6 | 1.98 | 1.59 | 2.46 |
|  | 12.95 | 23 | 20 | 87.0 | 58 | 22 | 37.9 | 2.29 | 1.59 | 3.30 |
|  | 13.35 | 41 | 28 | 68.3 | 40 | 14 | 35.0 | 1.95 | 1.22 | 3.13 |
|  | 13.45 | 46 | 31 | 67.4 | 35 | 11 | 31.4 | 2.14 | 1.26 | 3.64 |
|  | 13.65 | 50 | 34 | 68.0 | 31 | 8 | 25.8 | 2.64 | 1.41 | 4.93 |
|  | 13.85 | 56 | 36 | 64.3 | 25 | 6 | 24.0 | 2.68 | 1.30 | 5.53 |
|  | 14.15 | 62 | 39 | 62.9 | 19 | 3 | 15.8 | 3.98 | 1.39 | 11.45 |
|  | 14.35 | 66 | 39 | 59.1 | 15 | 3 | 20.0 | 2.95 | 1.05 | 8.29 |

Relative risk (RR) and 95% confidence intervals (CI) for developing anemia at month 1 (M1) among 120 initially non-anemic male patients (Hb ≥13.0 g/dL) and 81 initially non-anemic female patients (Hb ≥12.0 g/dL) treated with MDT/WHO. Patients were stratified by baseline hemoglobin values (M0) using incremental thresholds (0.1 g/dL), and RR values were calculated for each cutoff. Anemia was defined according to WHO criteria. Dashes (—) indicate null value.

Among patients with Hb≤14.0g/dL, the incidence of anemia was 71.0%, compared to 28.1% in those with Hb >14.0g/dL (RR: 2.53; 95% CI: 1.69-3.77). For the cutoff of Hb≤14.3g/dL, at M1, 59.1% of patients developed anemia, versus 27.6% in the group above this threshold (RR: 2.14; 95% CI: 1.38-3.32).

At the cutoff of Hb≤14.5g/dL, the incidence was 56.1%, compared to 23.8% in the higher group (RR: 2.36; 95% CI: 1.43-3.88). For Hb≤15.0g/dL, 51.4% of patients became anemic, versus 22.0% with Hb >15.0g/dL (RR: 2.34; 95% CI: 1.32-4.13).

In the full series of cutoff levels, RR values remained above 2.0 for all tested thresholds between 13.0 and 15.0g/dL. Incidence of anemia progressively decreased with increasing baseline Hb values, ranging from 100.0% (Hb≤13.0) to 51.4% (Hb≤15.0), while the corresponding incidence in higher Hb strata remained consistently between 22.0% and 38.7%. The full database table for the male patients stratification of anemia and RR values is available at S1 Table.

Table 7 also presents the RR of anemia at M1 among 81 initially non-anemic female patients, stratified by baseline Hb levels at M0. For patients with Hb≤13.0g/dL, 79.3% developed anemia at M1, compared to 36.5% in those with Hb >13.0g/dL (RR: 2.17; 95% CI: 1.35–3.47). At the Hb≤13.1g/dL cutoff, anemia incidence was 77.4%, versus 36.0% among those above this value (RR: 2.15; 95% CI: 1.35–3.44).

When using Hb≤13.2g/dL as a threshold, 70.3% of patients developed anemia compared to 36.4% in the higher group (RR: 1.93; 95% CI: 1.22-3.01). At the cutoff of Hb≤13.5g/dL, the anemia incidence was 68.0%, compared to 25.8% above this level (RR: 2.64; 95% CI: 1.41-4.93). The same RR was observed at Hb≤13.6 and≤13.7g/dL, with incidence rates above 66% in both strata.

From Hb≤13.8g/dL onward, the incidence of anemia dropped below 65%, with RR values still above 2.0. At Hb≤14.0g/dL, the incidence of anemia was 62.3%, compared to 20.0% in those with higher values (RR: 3.11; 95% CI: 1.27-7.65). Across all tested cutoff points, RR remained greater than 1.5 and above 2.0 in the majority of thresholds tested. The full database table for the female patients stratification of anemia and RR values is available at S1 Table.

### Evaluation of test performance as predictors of anemia

To determine predictive cutoffs for the development of anemia after dapsone initiation, ROC curve analysis was performed separately for male and female patients using M0 hematological parameters as predictors and anemia status at M1 as the outcome.

In male patients, Hb yielded an area under the curve (AUC) of 0.710 (95% CI: 0.608-0.807), with an optimal cutoff of 14.15g/dL (sensitivity=48.9%, specificity=86.3%; Youden Index=0.352). HCT and RBC count also demonstrated good predictive performance, with AUCs of 0.711 (95% CI: 0.611-0.801) and 0.599 (95% CI: 0.497-0.700), respectively. The best thresholds identified were 43.05% for HCT (sensitivity=68.1%, specificity=67.1%) and 4.88×10⁶/mm³ for RBC (sensitivity=59.6%, specificity=58.9%). The performance metrics are presented in Table 8, and the corresponding ROC curves are shown in Figure 5.

**Figure 5.**
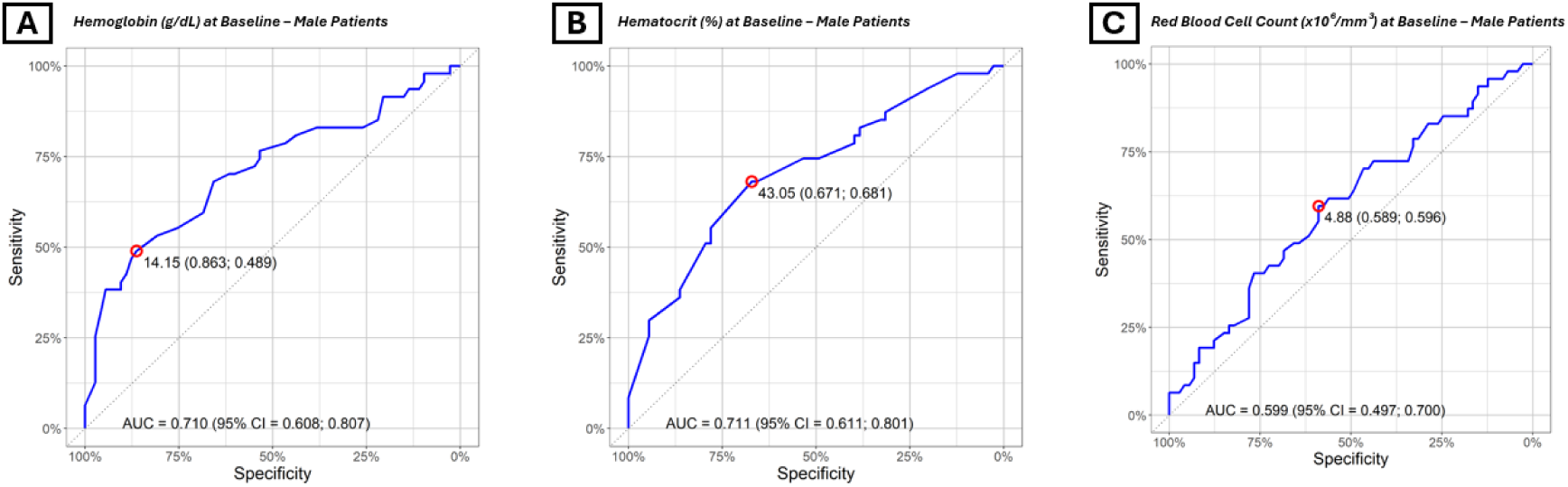
ROC curves for predicting anemia at month 1 in male patients based on baseline hematological parameters. (A) Hemoglobin (Hb) at baseline: AUC=0.710 (95% CI: 0.608-0.807), optimal cutoff=14,15g/dL (sensitivity=48.9%, specificity=86.3%). (B) Hematocrit (HCT): AUC=0.711 (95% CI: 0.611-0.801), optimal cutoff=43.05% (sensitivity=68.1%, specificity=67.1%). (C) Red blood cell (RBC) count: AUC=0.599 (95% CI: 0.497–0.700), optimal cutoff=4.88×10⁶/mm³ (sensitivity=59.6%, specificity=58.9%). All parameters showed good discriminative performance. Red open circles indicate the optimal cutoff points based on the Youden Index.

**Table 8.**
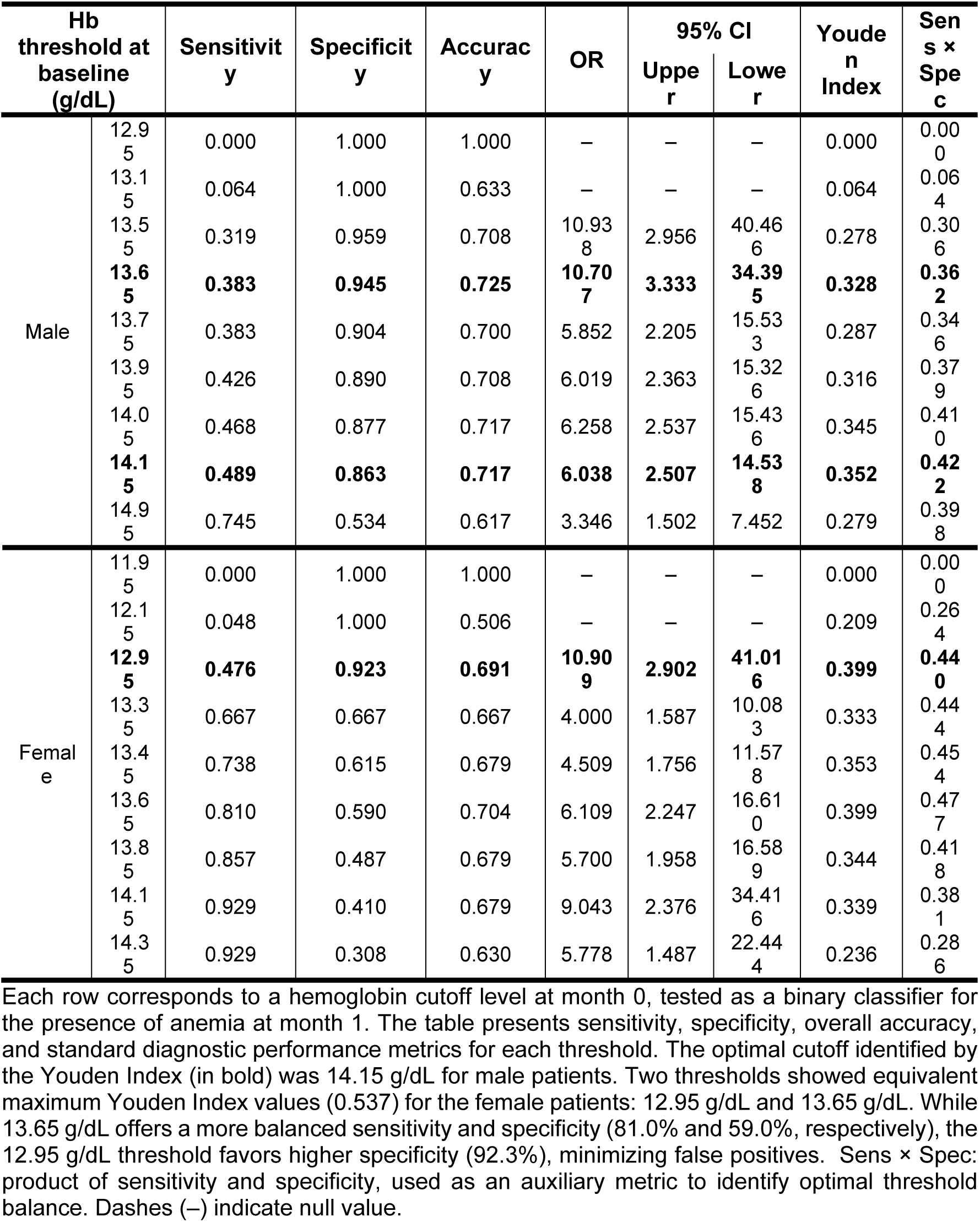
Performance metrics of ROC thresholds for predicting anemia at month 1 among male and female patients based on baseline hemoglobin.

In female patients, Hb had an AUC of 0.767 (95% CI: 0.657–0.863). Two cutoffs exhibited equal Youden Indices (0.399): 12.95g/dL (sensitivity=47.6%, specificity=92.3%) and 13.60g/dL (sensitivity=81.0%, specificity=59.0%). For HCT, the AUC was 0.561 (95% CI: 0.492–0.680), with an optimal threshold of 38.85% (sensitivity=83.3%, specificity=35.9%). The RBC count yielded an AUC of 0.590 (95% CI: 0.471–0.708), with a cutoff of 4.45×10⁶/mm³ (sensitivity=57.1%, specificity=66.7%). These results are summarized in Table 8 and illustrated in Figure 6. The full results for performance metrics and ROC curve analysis are available at S1 Table.

**Figure 6.**
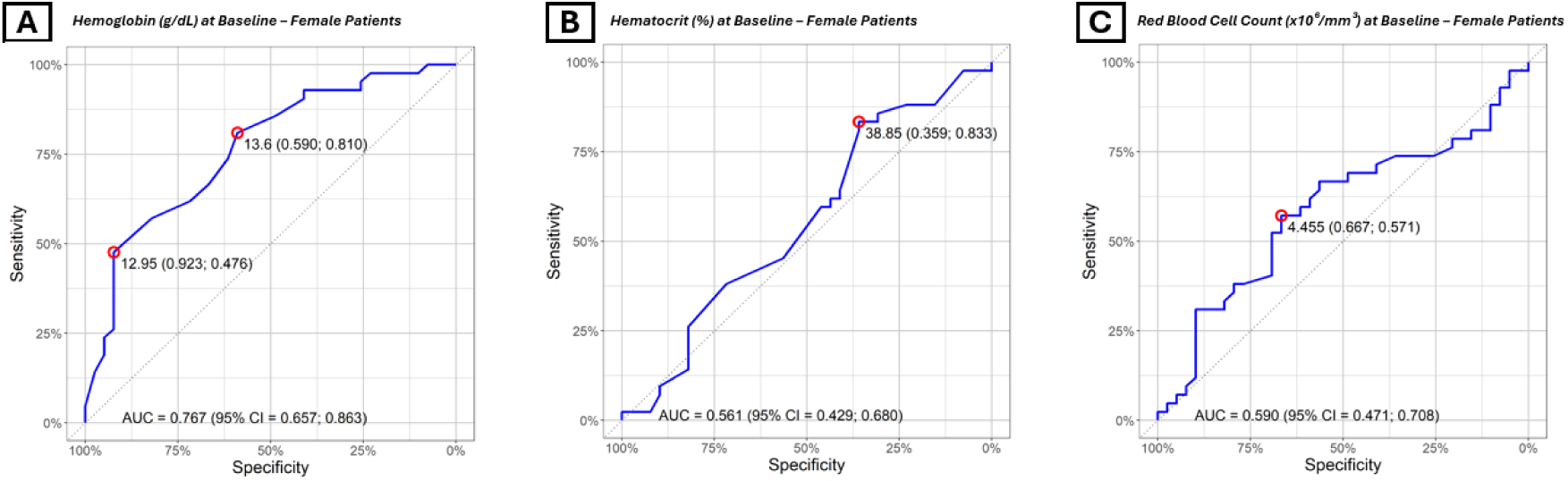
ROC curves for predicting anemia at M1 in female patients based on baseline hematological parameters. (A) Hemoglobin (Hb) at baseline: AUC=0.767 (95% CI: 0.657-0.863), with two equivalent optimal thresholds, 12.95g/dL (sensitivity=47.6%, specificity=92.3%) and 13.6g/dL (sensitivity=81.0%, specificity=59.0%). (B) Hematocrit (HCT): AUC=0.561 (95% CI: 0.492-0.680), optimal cutoff=38.85% (sensitivity=83.3%, specificity=35.9%). (C) Red blood cell (RBC) count: AUC=0.590 (95% CI: 0.471-0.708), optimal cutoff=4.45×10⁶/mm³ (sensitivity=57.1%, specificity=66.7%). Each curve evaluates the diagnostic accuracy of a baseline parameter for predicting anemia at M1. Red open circles indicate cutoff points with maximum Youden Index.

### Effect of MDT on the liver profile

No patient developed clinically significant hepatotoxicity or met laboratory criteria for acute liver injury during the course of treatment. While mild elevations in AST, ALT, and GGT were observed in some individuals, none required treatment interruption or regimen modification.

The descriptive evolution of hematological and biochemical parameters at months 0, 1, 3, 6, and 12 is provided in S2 Table.

Table 9 presents the Pearson correlation between percent change in Hb from M0 to M1 and the percent variation in other hematologic and biochemical parameters during the same period (n=258). Hb levels decreased from 13.68 (SD=1.69)g/dL at M0 to 12.27 (SD=1.89)g/dL at M1, corresponding to a mean reduction of 8.3%. The strongest positive correlation was observed between Hb and HCT (r=0.9441; R²=0.8914; p<0.0001), followed closely by RBC (r=0.9267; R²=0.8588; p<0.0001), indicating that reductions in Hb were consistently accompanied by proportional decreases in both RBC and HCT.

**Table 9.** Pearson correlation between percent change in Hb from M0 to M1 and changes in hematologic and biochemical parameters during the same period (n=258).

| Comparison | Pearson r | 95% CI | R<sup>2</sup> | p-value |
| --- | --- | --- | --- | --- |
| Hb vs. RBC | 0.9267 | 0.9073 to 0.9422 | 0.8588 | <0.0001 |
| Hb vs. HCT | 0.9441 | 0.9291 to 0.9560 | 0.8914 | <0.0001 |
| Hb vs. WBC | -0.1450 | -0.2624 to -0.0232 | 0.0210 | 0.0198 |
| Hb vs. PLT | -0.3337 | -0.4380 to -0.2206 | 0.1114 | <0.0001 |
| Hb vs. AST | -0.0600 | -0.1806 to 0.0628 | 0.0036 | 0.3389 |
| Hb vs. ALT | -0.0600 | -0.1806 to 0.0628 | 0.0036 | 0.3389 |
| Hb vs. ALP | -0.2536 | -0.3644 to -0.1357 | 0.0643 | <0.0001 |
| Hb vs. GGT | -0.2199 | -0.3330 to -0.1004 | 0.0483 | 0.0004 |

RBC: red blood cell count; HCT: hematocrit; WBC: white blood cell count; PLT: platelet count; AST: aspartate aminotransferase; ALT: alanine aminotransferase; ALP: alkaline phosphatase; GGT: gamma-glutamyl transferase. Percent change=((M1-M0)/M0) × 100

A weak negative correlation was found between Hb and platelet count (PLT) (r=−0.3337; R²=0.1114; p<0.0001), with a slight increase in mean PLT from 246.39±92.45 ×10³/mm³ at M0 to 246.40±105.93 ×10³/mm³ at M1.

Liver enzymes AST and ALT showed no significant correlation with Hb changes (r=−0.0600 for both; p=0.3389), and both displayed large interindividual variability at M1 (mean AST: 44.26±243.70 U/L; ALT: 44.26±243.70 U/L), likely influenced by outliers.

Weak negative correlations were observed for alkaline phosphatase (ALP; r=−0.2536; R²=0.0643; p<0.0001) and gamma-glutamyl transferase (GGT; r=−0.2199; R²=0.0483; p=0.0004), whose means increased from 206.48±96.87 to 230.88±317.21 U/L (ALP) and from 52.38±72.65 to 64.62±116.94 U/L (GGT), respectively, despite high variability.

## Discussion

The results provide robust evidence that dapsone induces a rapid and clinically significant decline in erythroid parameters, most notably Hb, within the first month of treatment, with disproportionate impact on female patients.

In a cohort study of 194 leprosy patients treated with MDT/WHO in Brazil between 1998 and 2003, hemolytic anemia occurred in 48 of them, with symptoms appearing predominantly within the first three months of therapy. Hb and HCT levels showed significant reductions, declining from 12.8g/dl to 10.3g/dl and from 38.5% to 31.5%, respectively, after dapsone initiation[24].

In our study, the early Hb reduction was accompanied by a sharp rise in anemia prevalence, particularly among individuals with lower baseline values. Substitution of dapsone after one month facilitated hematologic recovery, whereas continued exposure in the MDT/WHO group led to sustained anemia over the 12-month course. Notably, nearly 95% of patients who were anemic at M0 remained anemic at M1, and these patients had baseline Hb levels below the defined normal range by WHO, reinforcing the causal association between dapsone use and anemia persistence.

Modifications or discontinuations of therapeutic regimens due to dapsone-related adverse events have also been documented in other studies. In a retrospective Brazilian cohort of 194 patients receiving MDT/WHO, 85 individuals (44%) experienced adverse effects attributable to dapsone, leading to this drug discontinuation in 46 cases (54% of those affected)[25]. An Indian cohort study reported that 36 patients developed adverse reactions to MDT/WHO requiring treatment interruption or modification. Among them, 26 patients specifically required discontinuation or substitution of dapsone[26]. In the Brazilian Uniform-MDT trial, 24 patients (3.2% of the cohort) discontinued dapsone due to adverse effects and were switched to an alternative regimen. All affected patients developed at least mild anemia, and methemoglobinemia was identified in one case[27].

Sex-stratified analyses revealed divergent trajectories: male patients, despite higher baseline anemia percentage, showed more efficient recovery over time, while females exhibited a proportionally greater increase in anemia prevalence, even after dapsone withdrawal. These differences likely reflect how a combination of lower iron reserves[28] and hormonal factors[29] affect the erythropoietic capacity, and underscore the need for sex-specific monitoring protocols.

In the Uniform MDT/WHO clinical trial in Brazil, 201 out of the 753 patients (26.7%) presented anemia as an adverse effect. Female patients experienced significantly greater reductions in RBC, HCT, and Hb than males. Across all groups, Hb levels declined sharply after the first MDT/WHO dose, followed by gradual recovery, more pronounced in males[27].

To enhance clinical predictability, we conducted risk stratification using relative risk and ROC curve modeling. Anemia could occur even in patients initially classified as non-anemic, with progressively higher risk as baseline Hb declined. In men, the risk exceeded twofold below 14.0g/dL, while in women it became significant below 13.4g/dL and peaked below 12.4g/dL.

Tables 8 refined these insights by identifying optimal thresholds through ROC curve analysis. In men, the Hb cutoff of 14.15g/dL (AUC=0.710) offered specificity of 86.3% and sensitivity of 48.9%. In women, thresholds of 12.95g/dL and 13.6g/dL yielded identical Youden Indices, but 12.95g/dL achieved a better balance between sensitivity and specificity.

We defined gray zones as intermediate value ranges in which the risk–benefit ratio of using dapsone should be carefully evaluated. These ranges were determined based on the odds ratio (OR) and relative risk (RR), with the ROC curve cutoff corresponding to the upper limit of each gray zone. For men, the Hb gray zone ranged from 13.65g/dL (OR=10.71, RR=2.76) to 14.15g/dL (OR=6.04, RR=2.53). For women, the Hb values ranged from 12.95g/dL (OR=10.91, RR=2.29) to 13.6g/dL (OR=6.11, RR=2.64).

Importantly, HCT and RBC count also showed predictive performance. In men, the AUCs for HCT and RBC were 0.711 and 0.599, respectively, with optimal cutoffs of 43.05% and 4.88×10⁶/mm³. In women, HCT (AUC=0.561) and RBC (AUC=0.590) performed similarly, with cutoffs of 38.85% and 4.45×10⁶/mm³. These findings indicate that anemia risk is reflected across multiple erythroid indices and that HCT and RBC can serve as complementary screening tools, although Hb remains the main indicator of anemia.

For man. we defined the gray zone of HCT from 40.95% (OR=7.31. RR=2.4) to 43.05% (OR=4.35. RR=2.44) and RBC from 4.62×10⁶/mm³ (OR=2.23. RR=1.58) to 4.88×10⁶/mm³ (OR=2.11. RR=1.57). For women. we defined the gray zone of HCT from 36.5% (OR=3.41. RR=2.13) to 38.85% (OR=2.8. RR=1.75) and RBC from 4.2×10⁶/mm³ (OR=3.92. RR=1.68) to 4.45×10⁶/mm³ (OR=2.67. RR=1.58). These indices serve as supportive parameters during the diagnosis of leprosy, as the primary criterion for deciding whether to initiate dapsone therapy is the Hb level.

Baseline full blood count (such as Hb, HCT and RBC) and follow-up at 4 and 8 weeks after MDT/WHO initiation are essential for early detection of adverse effects. Patients and caregivers should be counseled on potential risks, warning signs, and appropriate actions. Proper training of healthcare providers is crucial to ensure effective patient education, monitoring, and timely management of anemia adverse events[30].

Taken together, these analyses support the definition of zones of contraindication and intermediate risk about the use of dapsone not only for Hb, but also for HCT and RBC count. Within the gray zones, decisions should be guided by nutritional status, comorbid conditions, reproductive age, and the capacity for close laboratory follow-up.

Corroborating these findings, Table 9 revealed strong positive correlations between changes in Hb and both HCT (r=0.9441) and RBC (r=0.9267), underscoring the internal consistency of the erythroid response. Conversely, liver transaminases (AST and ALT) showed no significant correlation, and other biochemical changes such as platelet count or alkaline phosphatase were modest and likely incidental.

In summary, this stratified data-driven approach allows for safer and more precise allocation of dapsone within multidrug regimens. It reinforces the principle of individualized medicine, anchored in objective laboratory thresholds rather than arbitrary eligibility. Implementation of pre-treatment screening using Hb, HCT, and RBC, interpreted by sex-specific cutoffs, can significantly enhance patient safety in endemic settings.

Furthermore, the observed difference in anemia persistence between groups not only corroborated the study hypothesis but also exceeded the estimated effect size used in the statistical power calculation. Whereas the study was powered to detect a 30% improvement in anemia resolution with dapsone substitution, the actual difference observed between M1 and M12 was 45.5 percentage points, favoring the SUBSTITUTIVE group. This magnitude of effect strengthens the clinical relevance of early dapsone discontinuation and supports the validity of the proposed cutoff thresholds.

The findings reinforce that anemia induced by dapsone was more severe and frequent in females, and its substitution after the first month (MDT/SUBSTITUTIVE) enabled partial recovery, though with residual anemia in a significant proportion, especially among women. The data also highlight that severe anemia was rare and transient, observed only in early time points within the MDT/SUBSTITUTIVE group.

Unfortunately, haptoglobin levels, reticulocyte counts, and RBC morphology were not available in this study, and these data could have aided in confirming the diagnosis of dapsone induced hemolytic anemia. As a retrospective analysis, patients in the HC FMRP–USP cohort did not have access to these laboratory investigations.

### Clinical recommendations

Based on the findings of this study, we propose the following clinical recommendations for the use of dapsone in leprosy treatment: Baseline Hb, HCT, and RBC count screening should be mandatory prior to initiating MDT/WHO regimens, with sex-specific interpretation of results.

#### For Male Patients

##### Absolute contraindication

Dapsone is formally contraindicated if any one of the following baseline criteria is met, due to a substantially elevated risk of developing anemia:

- Hb<13.65g/dL (RR of anemia at M1: 2.76[95% CI: 1.92-3.98])
- HCT<40.95%
- RBC<4.62×10⁶/mm³

##### Relative contraindication (individualized assessment)

Dapsone prescription requires criterious individualized assessment if any one of the following baseline criteria is met. Within this range, corresponding to the gray zone, it is imperative to consider factors such as nutritional status, comorbid conditions, and monitoring capacity:

- Hb<14.15g/dL (RR of anemia at M1: 2.53[95% CI: 1.68-3.80])
- HCT<43.05%
- RBC<4.88×10⁶/mm³

#### For Female Patients

##### Absolute contraindication

Dapsone is formally contraindicated if any one of the following baseline criteria is met, due to a substantially elevated risk of developing anemia:

- Hb<12.95g/dL (RR of anemia at M1: 2.29[95% CI: 1.59-3.30])
- HCT<36.5%
- RBC<4.2×10⁶/mm³

##### Relative contraindication (individualized assessment)

Dapsone prescription follows the same precautions mentioned before for relative contraindication for male patients:

- Hb<13.65g/dL (RR of anemia at M1: 2.64[95% CI: 1.41-4.93])
- HCT<38.85%
- RBC<4.455×10⁶/mm³

In resource-limited settings or in cases of anemia with conflicting markers, the combined use of Hb, HCT, and RBC thresholds increases diagnostic robustness and should guide regimen selection and follow-up testing. Alternative regimens containing ofloxacin or minocycline should be considered in at-risk individuals, as these may offer similar efficacy with lower hematologic toxicity.

Follow-up Hb testing is advised after one month of therapy, especially in patients near the established thresholds, to ensure early detection of treatment-related anemia.

In cases of anemia development during treatment, prompt substitution of dapsone should be pursued to allow for hematologic recovery and prevent long-term suppression.

These recommendations aim to enhance treatment safety without compromising efficacy and should be adapted according to regional resources and patient profiles.

## Conclusions

This study demonstrates that dapsone use in multidrug therapy for leprosy is associated with a significant and early decline in Hb levels, particularly in patients with lower baseline values. Through the use of ROC modeling and relative risk analysis, we identified clinically relevant and sex-specific thresholds based on Hb, HCT, and RBC count that predict the development of anemia during the first month of treatment.

These results support an individualized, multi-marker approach to dapsone prescription, expanding safety assessment beyond Hb alone. The combined use of Hb, HCT, and RBC cutoffs provides flexibility in clinical decision-making and allows safer treatment allocation, especially in settings with variable testing availability. Further research with larger and more diverse populations is warranted to validate the proposed thresholds and determine their applicability across different demographic and epidemiologic contexts. Future studies should also explore genetic, nutritional, and pharmacologic factors that may modulate individual susceptibility to dapsone-related anemia.

## Data Availability

The raw data supporting the conclusions of this article will be made available by the authors without undue reservation.

## Acknowledgments

We thank the team members and all patients who participated in this study.

## Author Contributions

CMS, MACF and FRL contributed to the conception and design of the study. CMS, GSA and FRL made substantial contributions to data acquisition, analysis and interpretation. FACC, HBL and MACF contributed to the clinical care of patients. JAW provided clinical data acquisition. FAMCP interpreted laboratory tests. CMS, GSA and FRL contributed to statistical analysis and data interpretation. MACF and FRL provided scientific guidance and advice. FRL and MACF approved the final submitted version and provided supervision and oversight of the study. All authors contributed to the interpretation of results and critical review of the manuscript.

